# The Immunology of Multisystem Inflammatory Syndrome in Children with COVID-19

**DOI:** 10.1101/2020.07.08.20148353

**Authors:** Camila Rosat Consiglio, Nicola Cotugno, Fabian Sardh, Christian Pou, Donato Amodio, Lucie Rodriguez, Ziyang Tan, Sonia Zicari, Alessandra Ruggiero, Giuseppe Rubens Pascucci, Veronica Santilli, Tessa Campbell, Yenan Bryceson, Daniel Eriksson, Jun Wang, Alessandra Marchesi, Tadepally Lakshmikanth, Andrea Campana, Alberto Villani, Paolo Rossi, the CACTUS study team, Nils Landegren, Paolo Palma, Petter Brodin

**Affiliations:** Science for life Laboratory, Dept. of Women’s and Children Health, Karolinska Institutet, Sweden; Research Unit of Congenital and Perinatal Infections, Bambino Gesù Children's Hospital, Rome, Italy; Chair of Pediatrics, Dept. of Systems Medicine, University of Rome “Tor Vergata”, Rome, Italy; Department of Medicine (Solna), Karolinska University Hospital, Karolinska Institutet, Sweden; Academic Department of Pediatrics, Bambino Gesù Children's Hospital, IRCCS, Rome, Italy; Center for Regenerative Medicine, Department of Medicine, Karolinska Institutet, Stockholm, Sweden; Department of Immunology, Genetics and Pathology, Uppsala University and Department of Clinical Genetics, Uppsala University Hospital, Uppsala, Sweden; Science for life Laboratory, Department of Medical Sciences, Uppsala University, Sweden; Pediatric Rheumatology, Karolinska University Hospital, Sweden

**Keywords:** SARS-CoV-2, COVID-19, Kawasaki disease, Multi-system inflammatory syndrome in children, MIS-C, Pediatric inflammatory multisystem syndrome temporally associated with SARS-CoV-2, PIMS-TS, Systems immunology, human immunology, autoantibodies, immunoglobulin

## Abstract

SARS-CoV-2 infection is typically very mild and often asymptomatic in children. A complication is the rare Multisystem Inflammatory Syndrome in Children (MIS-C) associated with COVID-19, presenting 4-6 weeks after infection as high fever, organ dysfunction and strongly elevated markers of inflammation. The pathogenesis is unclear but has overlapping features with Kawasaki disease suggestive of vasculitis and a likely autoimmune etiology. We apply systems-level analyses of blood immune cells, cytokines and autoantibodies in healthy children, children with Kawasaki disease enrolled prior to COVID-19, children infected with SARS-CoV-2 and children presenting with MIS-C. We find that the inflammatory response in MIS-C differs from the cytokine storm of severe acute COVID-19, shares several features with Kawasaki disease, but also differs from this condition with respect to T-cell subsets, IL-17A and biomarkers associated with arterial damage. Finally, autoantibody profiling suggests multiple autoantibodies that could be involved in the pathogenesis of MIS-C.

**HIGHLIGHTS:** Hyperinflammation in MIS-C differs from that of acute COVID-19

T-cell subsets discriminate Kawasaki disease patients from MIS-C

IL-17A drives Kawasaki, but not MIS-C hyperinflammation.

Global autoantibodies profiling indicate possibly pathogenic autoantibodies

## INTRODUCTION

The severe acute respiratory syndrome coronavirus 2 (SARS-CoV-2), first emerged in Wuhan in December 2019 (Huang et al., 2020) and then spread rapidly to a number of countries and in particular to Europe and the northern regions of Italy in the early weeks of February 2020. The first reports from China showed that children presented with milder symptoms as compared to adults infected by SARS-CoV-2 (Lu et al., 2020). Reasons for this have not been established, but several theories have been discussed, involving immune system differences such as thymic function, cross-reactive immunity to common cold coronaviruses, as well as differences in the expression of the viral entry receptor ACE2, as well as a better overall health status among children as compared to the elderly (Brodin, 2020). The mild COVID-19 in children has been confirmed also in areas with high disease prevalence such as northern Italy (Parri et al., 2020). The view that COVID-19 disease course is always mild in children is now challenged by recent reports of children presenting with a rare, but very severe hyperinflammatory syndrome in the United Kingdom (Riphagen et al., 2020; Whittaker et al., 2020), Italy (Verdoni et al., 2020), Spain (Moraleda et al., 2020) and New York City (Cheung et al., 2020). In these case series, children present with high fevers, and a variable number of symptoms previously associated with Kawasaki disease such as conjunctivitis, lymphadenopathy, mucocutaneous rash and coronary artery dilation and in the most severe cases cardiovascular shock, encephalitis and multiple organ failure.

Kawasaki disease is a vasculitis affecting medium sized arteries with highest incidence in children younger than 5 years old and it is the leading cause of acquired heart disease in developed countries where streptococcal infections are commonly treated, and rheumatic fever is rare (Shulman and Rowley, 2015). Kawasaki disease has a particularly high incidence in children with East Asian ancestry and was first described in Japan. The dominating theory for the pathophysiology of Kawasaki disease involves the production of self-reactive antibodies during an acute immune response to a viral infection, probably at mucosal surfaces and focused around IgA-producing plasma cells. Such cells have also been found within the arterial wall in specimens from children with Kawasaki disease (Rowley et al., 2008). Neutrophils infiltrate the arterial wall and a necrotizing arteritis develops leading to the destruction of the connective tissue and arterial dilation in severe cases (Shulman and Rowley, 2015). Efforts to define the pathogenesis of Kawasaki have also revealed an imbalance of IL-17-producing T-cells and regulatory T-cells during the acute phase of this disease (Jia et al., 2010).

Given the severity of the Multisystem Inflammatory Syndrome in children (MIS-C) associated with COVID-19, and the uncertain development of the ongoing pandemic, there is an urgent need to understand the pathogenesis of MIS-C, its similarities and differences with Kawasaki disease, such that optimal treatment strategies can be devised. Here we have performed a systems-level analysis of immune cells, cytokines and antibodies in the blood of children presenting with MIS-C as compared to children with mild SARS-CoV-2 infection, children with Kawasaki disease and healthy children enrolled prior to the COVID-19 pandemic (**Figure 1**). We reveal several details of the hyperinflammatory state in children with MIS-C, contrasting with both the hyperinflammation seen in adults with acute SARS-CoV-2 infection, as well as that of children with Kawasaki disease. We define key cytokine mediators, assess serological responses to SARS-CoV-2 and other viruses and we identify putative autoantibody targets possibly involved in the pathogenesis of this new disease.

**Figure 1.**
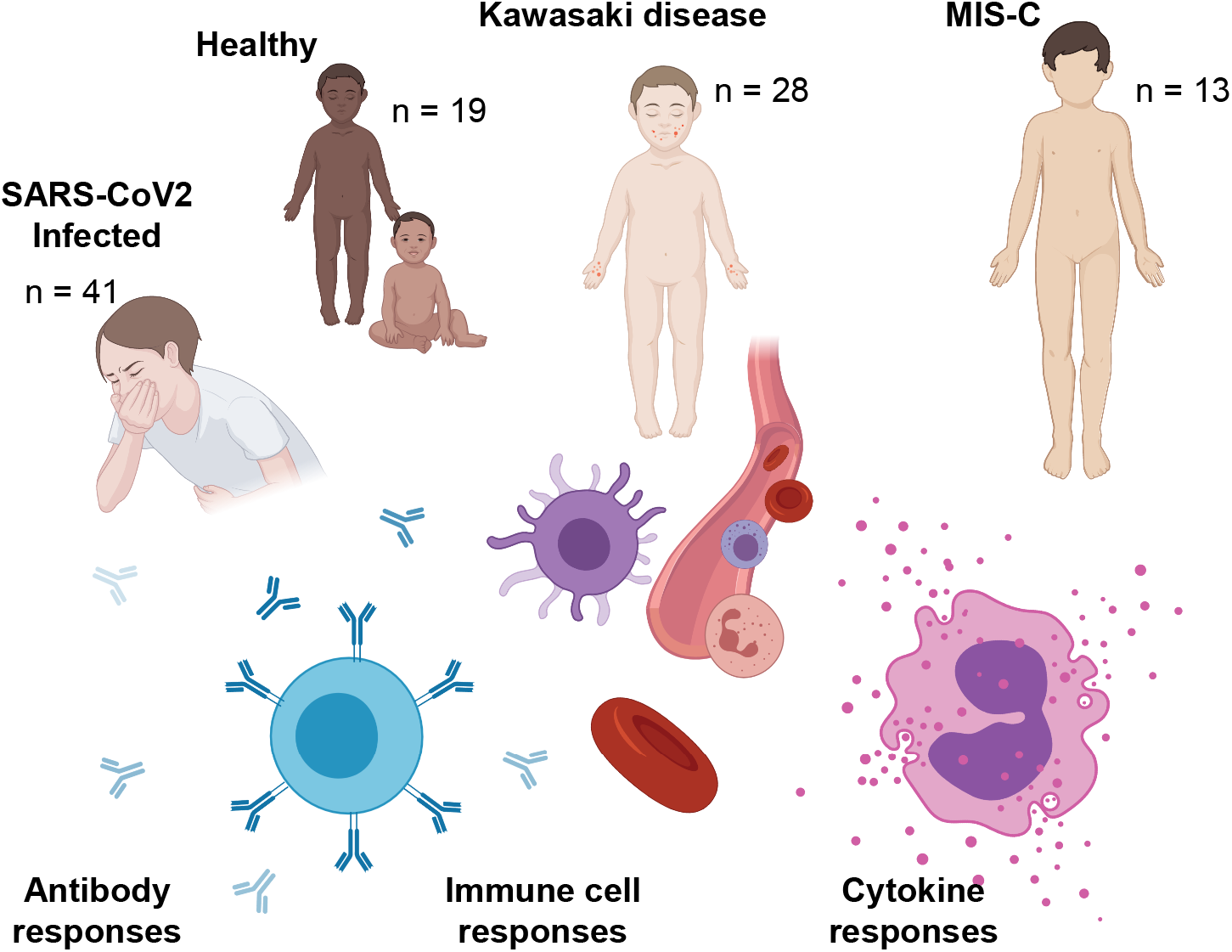
Systems-level analyses in children with COVID-19, Kawasaki disease and MIS-C

## RESULTS

### Children with SARS-CoV-2, MIS-C and Kawasaki disease

We enrolled 41 children with acute SARS-CoV-2 infection in Rome, Italy, all with mild disease, and we denote these as CoV2+ children throughout this manuscript (**Table 1**). We also enrolled three children presenting with MIS-C in Rome and 10 children presenting with MIS-C in Stockholm, Sweden. We compare these CoV2+ and MIS-C children with 28 children presenting with Kawasaki disease prior to the COVID-19 pandemic from March 2017 to May 2019. The children with MIS-C were significantly older than children with Kawasaki disease (**Table 1**), in line with previous reports (Whittaker et al., 2020). No significant sex-differences were found among the groups of children. Both MIS-C and CoV2+ children presented with lower white blood cell counts (WBC) as compared to patients with Kawasaki disease and healthy children (**Table 1**). Lymphopenia is a hallmark of COVID-19 and was more pronounced in MIS-C than in children with mild SARS-CoV-2 infection, and also more pronounced than in children with Kawasaki disease (**Table 1**). MIS-C patients also had markedly higher CRP and ferritin levels and lower platelet counts as compared to both Kawasaki disease and CoV2+ children (**Table 1**). These observations are in line with recent clinical reports (Whittaker et al., 2020), suggesting that our cohort is representative of patients with MIS-C, Kawasaki disease and children with mild SARS-CoV-2-infection.

**Table 1.**
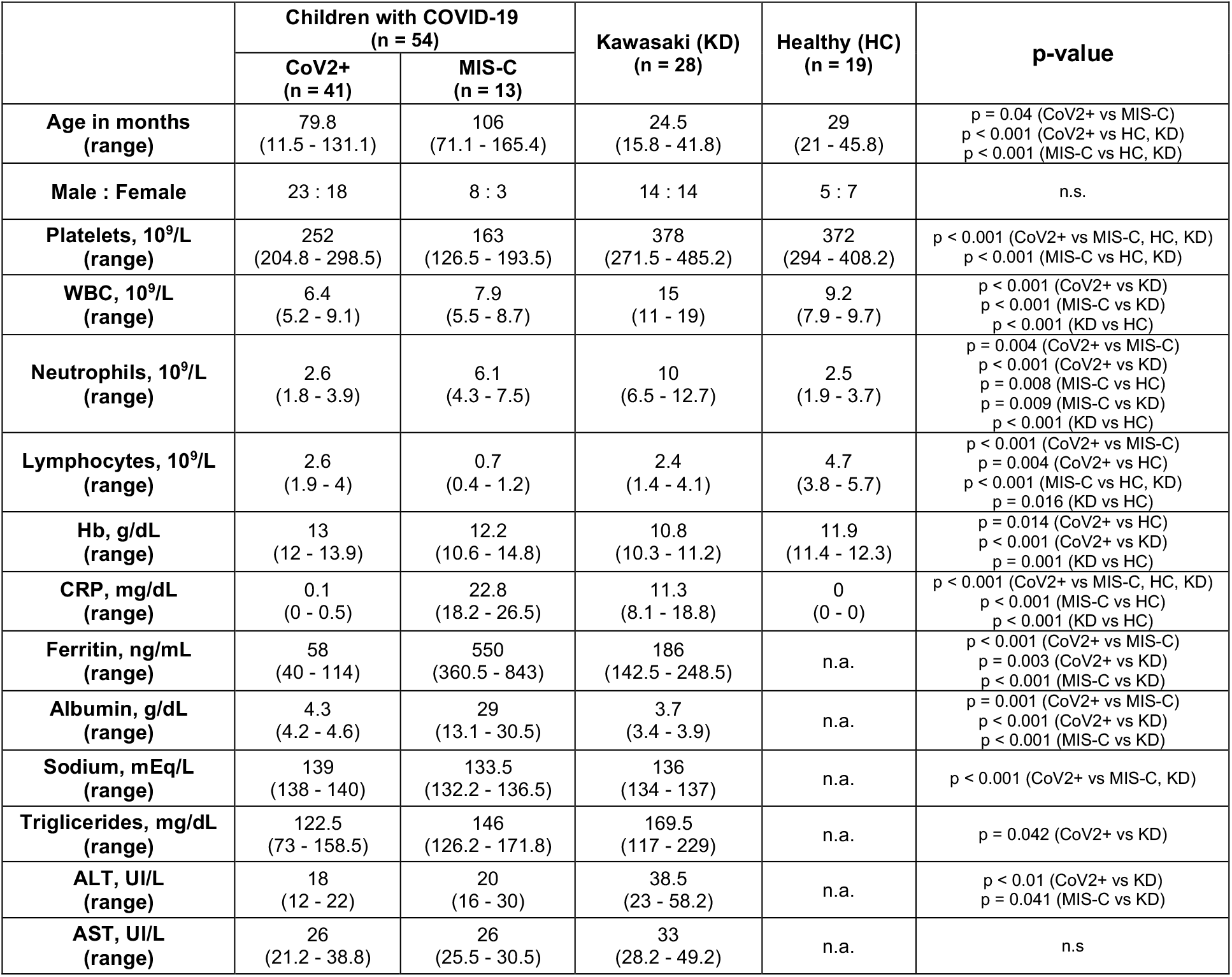
Clinical parameters presented as median, 25^th^ - 75^th^ centiles. T-test (parametric) and Mann-Whitney test (non-parametric) were used to compare mean values across groups of children.

### Hyperinflammation during MIS-C differs from that of severe, acute COVID-19

In adults and the elderly, the main cause of severe COVID-19 disease and death is uncontrolled immune activation, hyperinflammation and immunopathology (Vardhana and Wolchok, 2020). This life-threatening response requires urgent management and the immunological aspects of this process is under intense investigation (Kuri-Cervantes et al., 2020; Mathew et al., 2020; Rodriguez et al., 2020). A number of clinical trials are ongoing to test immunomodulatory strategies that can calm the cytokine storm in severe COVID-19. Since MIS-C is also a hyperinflammatory condition associated with COVID-19, but with a delayed presentation, we wondered whether this hyperinflammatory state is similar to that seen in adults with severe COVID-19 disease and whether similar immunomodulatory strategies should be considered in both conditions. We measured 180 plasma proteins involved in immune response and inflammation in serum samples from children with mild MIS-C and Kawasaki disease and compared these to the cytokine profiles recently reported in adults with severe acute COVID-19 and hyperinflammation (Rodriguez et al., 2020). After filtering out proteins with >30% measurements below the threshold of detection, we performed principal component analysis (PCA) including 120 unique proteins (**Figure 2A, Supplementary Table 1**). Adults with acute COVID-19, both patients in the Intensive Care Unit (ICU) and floor (non-ICU), but all severely ill, had very different cytokine profiles from those seen in children with either MIS-C or Kawasaki disease (**Figure 2A**). The main contributing features explaining this difference included IL-8 (**Figure 2B**), a chemokine recently shown to be associated with lymphopenia in severe COVID-19 cases (Zhang et al., 2020). IL-7 was also higher in acute COVID-19 hyperinflammation than in MIS-C and Kawasaki disease, IL-7 is a cytokine involved in T-cell maintenance and associated with lymphocyte counts (**Figure 2B**). The MIS-C and Kawasaki hyperinflammatory states partially overlapped and both differed from adults with acute COVID-19 hyperinflammation (**Figure 2A**).

**Figure 2.**
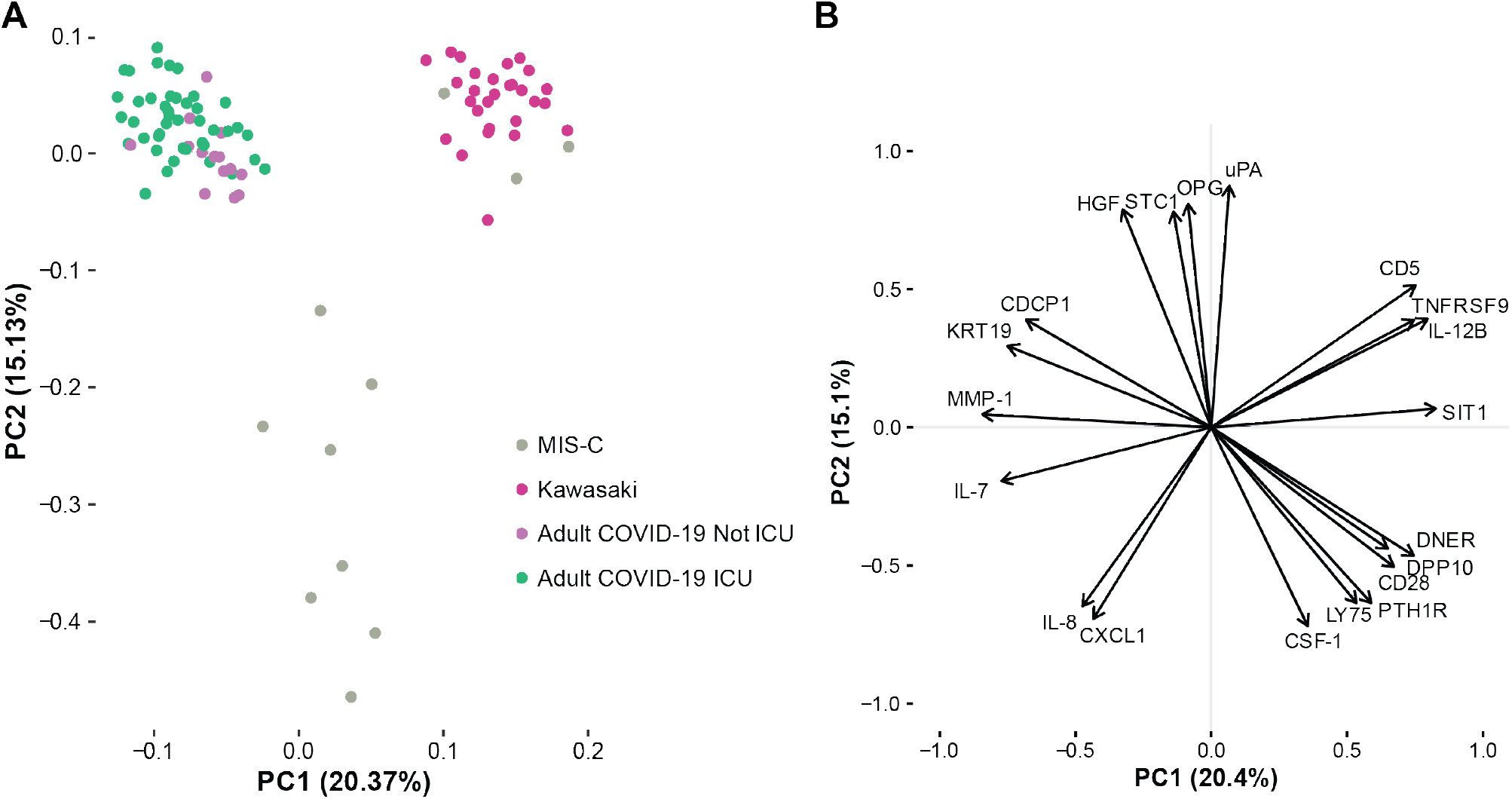
MIS-C hyperinflammation differs from severe acute COVID-19 hyperinflammation. **(A)** Principal components 1 and 2 show variation in cytokine profiles among adult COVID-19 patients with severe disease treated in intensive care units, ICU or not and children with MIS-C or Kawasaki disease. n=97 samples included and 112 unique proteins included in the analysis. **(B)** top 20 proteins mostly contributing to the PCs 1-2.

### Differences in T-cell subsets in MIS-C and Kawasaki disease

To better understand the hyperinflammation in MIS-C and Kawasaki disease we assessed peripheral blood mononuclear cell (PBMC) phenotypes by flow cytometry. Samples were collected at hospital admission, prior to treatment and reflect cellular states during the hyperinflammatory immune response. We found differences in the distributions of subpopulations of CD4^+^ T-cells as defined by the expression of CD45RO and CD27, and the frequency of T-follicular helper cells (T_FH_) expressing the chemokine receptor CXCR5 (). Total T-cell frequencies were lower in both types of hyperinflammatory patients, MIS-C and Kawasaki disease as compared to healthy children (**Figure 3A**). Within the CD4^+^ T-cell compartment, MIS-C patients and children with mild SARS-CoV-2+ infection had similar subset distributions, indicating that differences seen in relation to healthy children could be related to the SARS-CoV-2 infection itself (**Figure 3C-E**). Both SARS-CoV-2 experienced groups of patients had higher abundances of central memory (CM) and effector memory (EM) CD4^+^ T-cells, but fewer naïve CD4^+^ T-cells as compared to Kawasaki disease patients (**Figure 3C-E**). Follicular helper T-cells, important players in germinal center reactions and supporters of B-cell responses were reduced in SARS-CoV-2 experienced children, both with and without MIS-C, but not in Kawasaki disease patients (**Figure 3F**). CD57 marks terminally differentiated effector CD4^+^ T-cells and these have been shown to be reduced in adult patients with severe acute COVID-19 and acute respiratory distress syndrome (Anft et al., 2020). In children with mild COVID-19 and children with MIS-C we find higher levels of these terminally differentiated cells when comparing to Kawasaki disease patients and healthy children (**Figure 3G**). We find that CD4^-^ T-cells (mostly CD8+ T-cells) were significantly lower in MIS-C children as compared to children with mild SARS-CoV-2 infection (**Supplementary Figure 1A-D**). These results further emphasize that the hyperinflammation seen in acute, severe COVID-19 in adults, differs from that seen in MIS-C and also indicates some specific differences between immune cell responses in MIS-C and patients with Kawasaki disease occurring prior to the COVID-19 pandemic.

**Figure 3.**
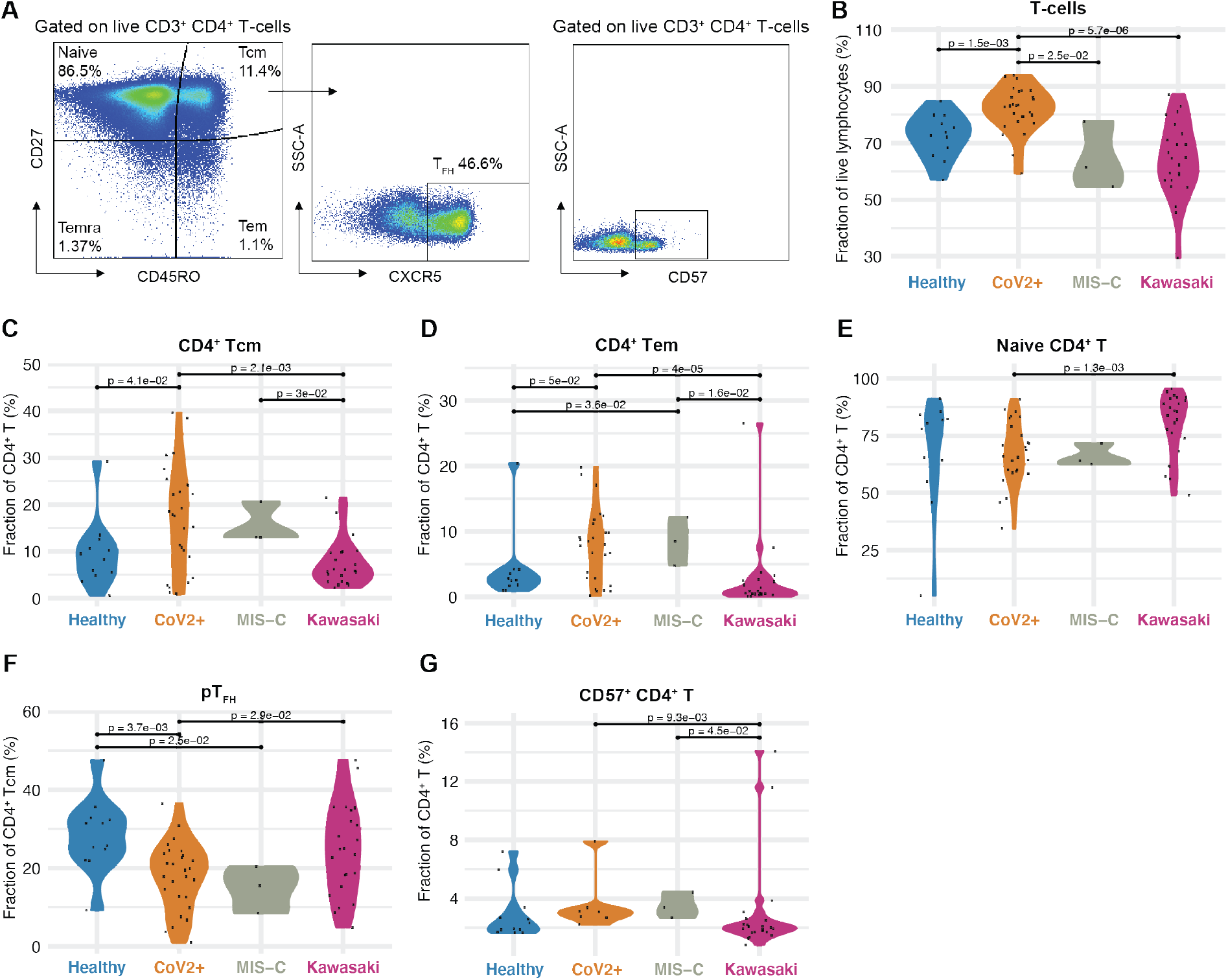
CD4 T-cell subsets in COVID-19 and MIS-C. **(A)** Gating schema to identify CD4^+^ T-ceii subsets from peripheral mononuclear cells. **(B)** CD4^+^ T-cells as a fraction of lymphocytes, **(C-G)** fraction of CD4^+^ T-cell subsets (%) in the indicated | group. Black lines indicate statistical tests and p-values across indicated populations.

### Unraveling the cytokine storm in MIS-C and Kawasaki disease

To further investigate the hyperinflammatory immune states in MIS-C and Kawasaki disease patients, we performed Olink assays on plasma samples from 11 children with MIS-C and 28 children with Kawasaki disease. We measured 180 unique proteins using two Olink panels **(Key Resource Table)** and filtered out proteins with >30% measurements below the threshold of detection, resulting in 125 plasma proteins used for PCA **(Figure 4A, Supplementary Table** 2).

We find that healthy children and children infected with SARS-CoV-2, without MIS-C, mostly overlapped, illustrating the mild infection and low-grade inflammatory response in these CoV2+ children (**Figure 4A**). As shown by the blown up PC2 vs. 3 plot, MIS-C hyperinflammatory states differ from that of Kawasaki disease (**Figure 4A**). PC2 best discriminated these hyperinflammatory states in MIS-C and Kawasaki disease and we focused on the contributing features (loadings) of PC2 (**Figure 4B**). We find that elevated IL-6, IL-17A, CXCL10 contributed the most to the cytokine storm (**Figure 4C**). IL-17A is important in Kawasaki disease (Jia et al., 2010), but was significantly lower in MIS-C patients, indicating a difference in the underlying immunopathology (**Figure 4C**). In contrast, the top negative contributors were adenosine deaminase (ADA), stem cell factor (SCF) and TWEAK, a negative regulator of IFNg and Th1-type immune response (Maecker et al., 2005). As a regulator of angiogenesis, TWEAK is also an interesting finding given the vasculitis component of Kawasaki disease and probably also MIS-C (**Figure 4D**).

**Figure 4.**
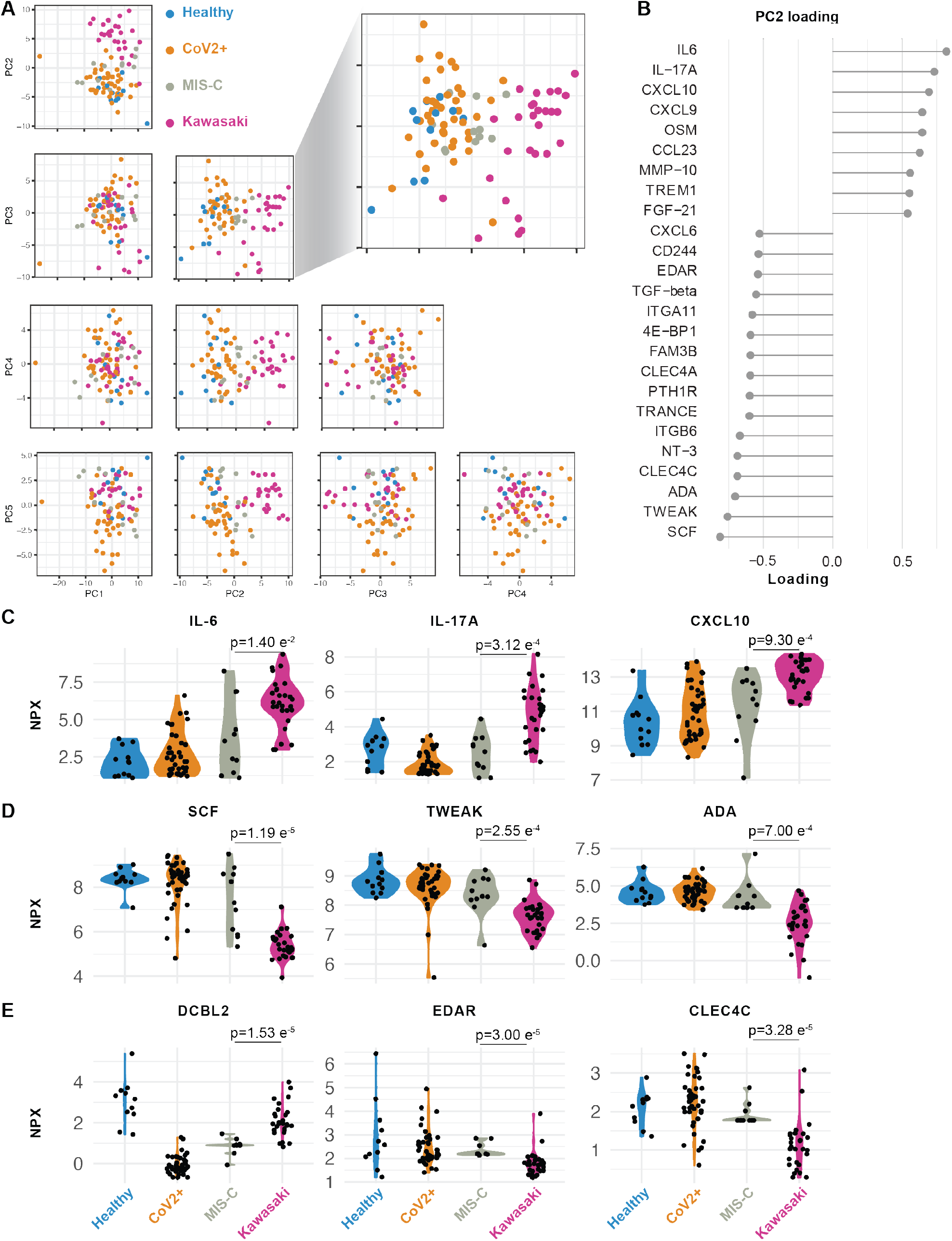
Cytokine profiles in MIS-C and Kawasaki disease. **(A)** Principal component analysis of top 5 components explaing 58.3% of the variance in concentrations of 133 plasma proteins. PC2 vs 3 separate children with Kawasaki (n=28) from MIS-C (n=11), healthy children (n=12) and SARS-CoV2+ children without hyperinflammation (n=41). **(B)** Proteins explaining (highest loading for) PC2. **(C-D)** Raw (NPX) values for top proteins (loading) in PC2. **(E)** Raw (NPX) values of three proteins significantly different between Kawasaki and MIS-C, p-values for mean comparisons between MIS-C and Kawasaki disease children.

We also identified additional plasma proteins distinguishing MIS-C from Kawasaki. Apart from IL-17A, DCBLD2, also called ESDN, was more elevated in Kawasaki disease than MIS-C. This protein has been shown to be secreted by endothelial cells attacked by allogeneic immune cells after heart transplantation (Sadeghi et al., 2007) and in balloon dilated carotid arteries (Kobuke et al., 2001) suggestive of more pronounced arterial damage in Kawasaki than in MIS-C. Collectively, these findings indicate more arterial involvement in Kawasaki than MIS-C, and also a IL-17A driven cytokine storm in Kawasaki disease but not in MIS-C. IL-17-blocking agents are used in psoriasis and other diseases and could be of interest for future clinincal trials also in these patients. Moreover, other key proteins differing among these groups is MMP-1 and MMP-10 (**Supplementary Figure 2A**), proteins involved in arterial disease (Martinez-Aguilar et al., 2015), which is intriguing given the likely arterial inflammation component of both MIS-C and Kawasaki disease.

### MIS-C immune profiles intersect mild COVID-19 and Kawasaki disease and change upon immunomodulation

To understand the interaction among different immune system components and their coregulation, we applied Multiomics Factor Analysis (MOFA) (Argelaguet et al., 2019) to integrate T-cell subset frequencies and plasma protein concentrations from MIS-C patients, Cov2+ children and children with Kawasaki disease. We identified 10 latent factors that explain the combined variance across cell and protein datasets (**Supplementary Figure 1D-E**). The first factor separated Kawasaki disease samples from both groups of SARS-CoV-2 infected children (MIS-C and CoV2+) (**Supplementary Figure 1F**). Conversely, factor 5 discriminated the extremes of Kawasaki vs. mild SARS-CoV-2+ infections and placed MIS-C samples in between these polar opposites (**Supplementary Figure 1G-H**), indicating that the immunopathology in MIS-C shares features with both acute SARS-CoV-2 infection and the postinfectious response associated with Kawasaki disease.

MIS-C patients are treated with strong immunomodulatory agents. The treatment regimens for the 13 MIS-C patients in our cohor are shown in **Figure 5A**. Most patients were treated by high dose steroids in combinations with intravenous immunoglobulins (IVIG) to override the effect of autoantibodies and stimulate inhibitory Fc-receptors (**Figure 5A**). A number of children were also given recombinant IL1RA (Anakinra) to block IL-1a/b mediated inflammation (**Figure 5A**). We assessed the cytokine changes upon treatment and found that TNFB, ITGA11 and CCL25 levels decreased, while HEXIM1, PSP1 and CXCL10 levels increased in response to treatment in the seven MIS-C patients with available pre/post samples (**Figure 5B-C**). These results indicate that there is a particular cytokine profile associated with MIS-C, that differs from Kawasaki disease hyperinflammation and that changes in response to immunomodulatory treatment.

**Figure 5.**
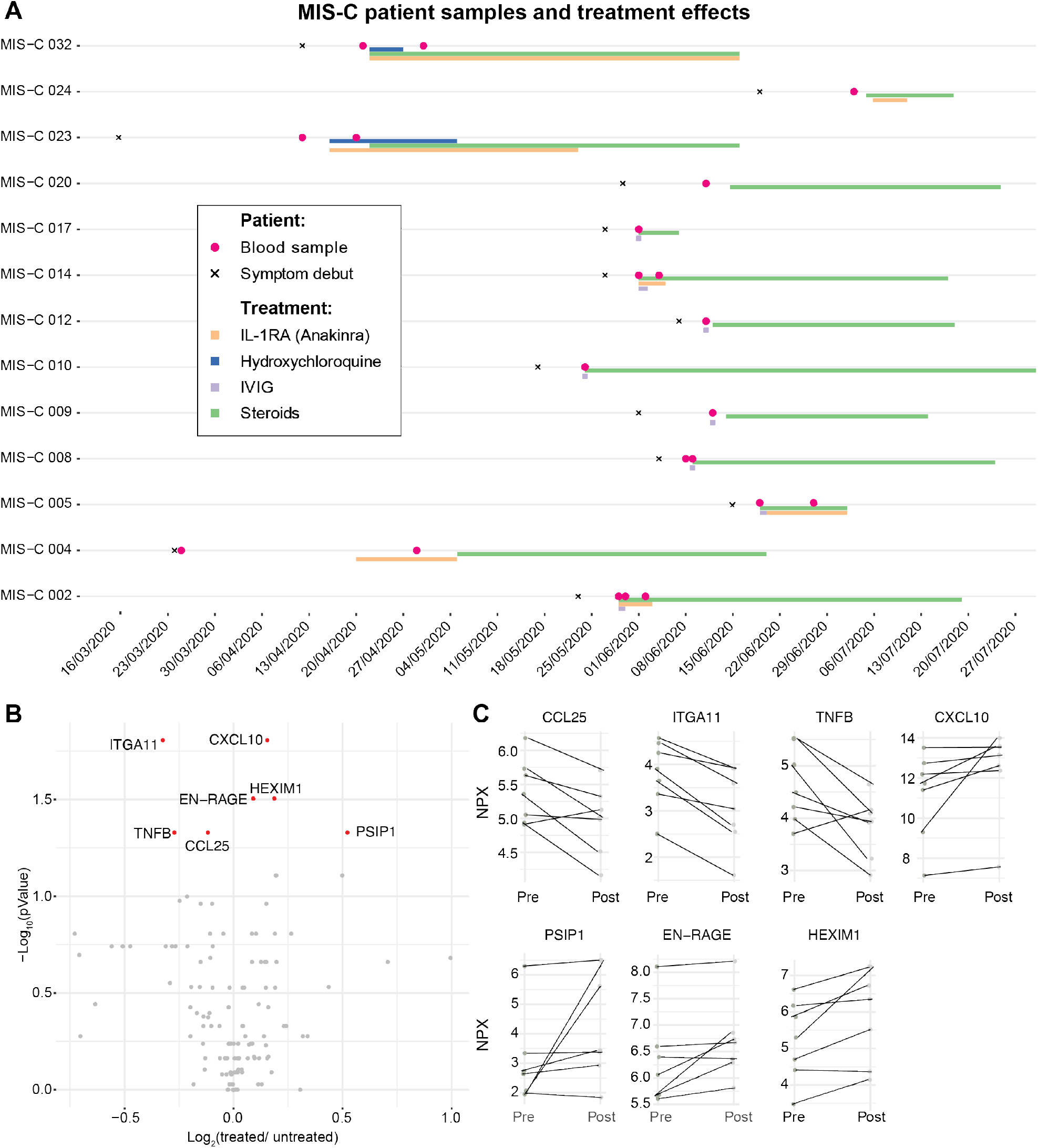
Immunomodulatory treatment in patients with MIS-C. **(A)** Timelines for individual MIS-C patients (n=13) indicating date of symptom debut, immunomodlatory treatment regimen, and date of blood sampling. Two patients lacking clinical treatment data were blood sampled prior to treatment. **(B)** Volcano plot showing fold-change (post/pre treatment) v.s. p-value for 7 patients with multiple samples. **(C)** Top markers of treatment response in MIS-C patients fold-change pre/post treatment (p-value <0.05, FDR 1%).

### Serologic responses and pre-existing humoral immunity in MIS-C

Given that immunopathology of Kawasaki disease and possibly also MIS-C involving autoantibodies, we decided to test IgG responses to the SARS-CoV-2 virus in these patients. We measured IgG binding of the receptor-binding domain (RBD) of the Spike protein from SARS-CoV-2. We found that the majority of children in our cohort testing positive for infection by PCR also seroconverted (**Figure 6A**). We also found that 3 out of 4 tested MIS-C patients seroconverted and had comparable levels of SARS-CoV-2 IgG antibodies to the spike protein as the children with mild SARS-CoV-2 infection and no MIS-C (**Figure 6A**). This result is in line with data from other groups studying antibody responses in MIS-C patients (Gruber et al., 2020; Rostad et al., 2020). A possible hypothesis for why some children develop MIS-C is that prior immunity to other viruses could modulate their responses to SARS-CoV-2 infection and give rise to hyperinflammation either by antibody-mediated enhancement or other mechanisms (Tetro, 2020). To assess this, we applied VirScan, a phage display method for testing IgG binding of 93,904 epitopes from 206 different viruses able to infect human cells (Xu et al., 2015). We found that the children in our cohort had IgG antibodies to common viruses such as respiratory syncytial virus (RSV) and rhinovirus as well as viruses of the herpesvirus family (**Figure 6B**). This is in line with our previous results (Pou et al., 2019). The common cold coronaviruses are particularly interesting as possible modulators of immune responses to SARS-CoV-2 given their similarities. We focused on IgG antibodies to these viruses in MIS-C, CoV2+, Healthy and Kawasaki-disease children. We found that IgG antibodies to human coronavirus HKU1 was commonly observed, as were antibodies to betacoronavirus 1, but the MIS-C patients were the only ones lacking antibodies to either of these common coronaviruses (**Figure 6C**). The relevance of this difference remains to be determined; this could either reflect the fact that the MIS-C patients are older than the other groups of children analyzed, but it is also possible that the lack of IgG antibodies to common coronaviruses modulates the immune response to SARS-CoV-2 infection and plays a role in the pathogensis of MIS-C.

**Figure 6.**
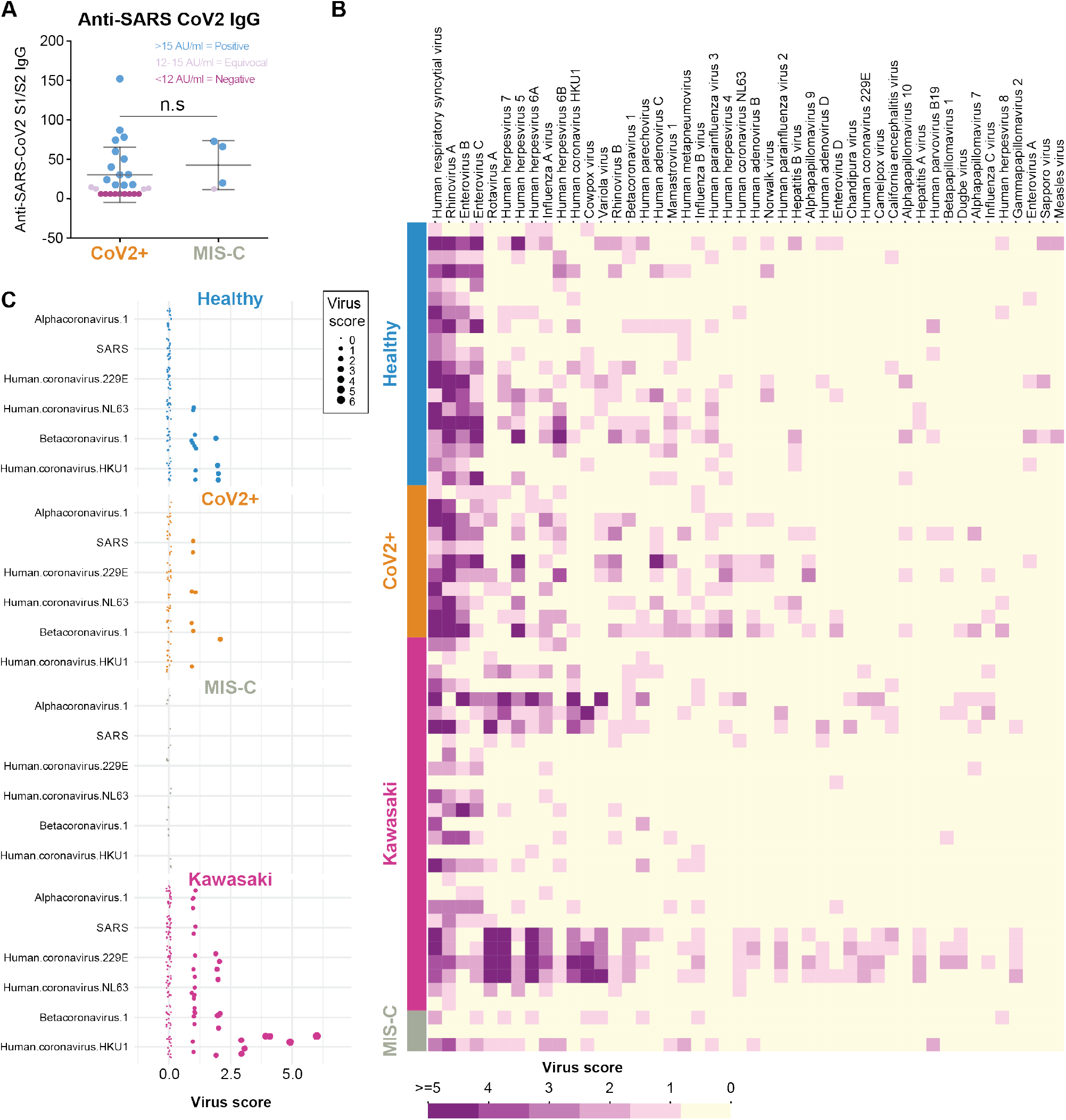
Serological responses and prior immunity in Kawasaki and MIS-C. **(A)** Anti-SARS-CoV2 S1/S2 IgG antibodies in patients with mild infection (n=26) and MIS-C (n=4). **(B)** VirScan analysis of IgG responses to 93,904 epitopes of 206 viruses in Healthy (n=19), CoV2 infected (n=11), Kawasaki disease (n=27) and children with MIS-C (n=3). Virus scores is a function of the number of targeted epitopes per virus and the heatmap shows the top 46 viruses with top average virus scores. **(D)** The virus scores of coronavirses available in the VirScan library, shown across individual children divided by disease groups.

### Proteome array profiling reveals potentially pathogenic autoantibodies in MIS-C

Several studies have proposed autoantibodies as an element of the immunopathology of Kawasaki disease (Sakurai, 2019). An external trigger, likely in the form of a virus is indicated by epidemics of Kawasaki disease, and direct evidence of antiviral antibody responses, particularly targeting viral particles in inclusion bodies in the respiratory epithelium have been reported (Rowley et al., 2008). Autoantibodies targeting endothelial cells can activate IL-6 production by endothelial cells in vitro (GRUNEBAUM et al., 2002), but a possible role of autoantibodies in MIS-C is currently unknown. To search for such autoantibodies in MIS-C, we screened serum samples from children with MIS-C (n=12), Kawasaki disease (n=28), SARS-CoV-2+ infection (n=5) and healthy control children (n=11) against 9341 human protein antigens from 7669 unique proteins using protein arrays (Zhu et al., 2001) (ProtoArray v5.1, PAH05251020, ThermoFisher, Waltham, Mass). As expected, the autoantibody signals were low for the vast majority of antigenic targets (Landegren et al., 2016) (**Figure 7A**). We ranked autoantibody targets using fold-change calculations between the MIS-C group and each of the other groups of samples, and looked for enriched Gene Ontology (GO)-terms among the targets using gene set enrichment analysis (GSEA) (Subramanian et al., 2005). There were 26 GO-terms that were enriched in MIS-C samples when compared to all other groups (**Figure 7B**), and these involved lymphocyte activation processes, phosphorylation signaling pathways and heart development (**Figure 7C**). The latter was interesting given that myocarditis and impaired cardiac function are hallmarks of MIS-C clinical presentation. We studied individual autoantibody targets within this GO-term (**Figure 7D**), and identified Endoglin, a glycoprotein expressed by endothelial cells and necessary for structural integrity of arteries to be differentially regulated among groups of samples (**Figure 7E**). Previous reports have found that when Endoglin is disrupted it causes the disease Hereditary hemorrhagic telangiectasia, a disease characterized by multisystemic vascular dysplasia (McAllister et al., 1994). Several, but not all of the MIS-C patients had elevated levels of autoantibodies targeting Endoglin higher than levels in healthy controls and a subset of Kawasaki disease patients also had autoantibodies to Endoglin (**Figure 7E**). Endoglin protein expression is seen predominantly in the vascular endothelium with the heart muscle having the highest mRNA expression of all tissues (Uhlen et al., 2015). Among immune cells, mainly monocytes express Endoglin (Uhlen et al., 2019). We also measured plasma levels of Endoglin, but in contrast to our expectation, the levels where elevated in Kawasaki and MIS-C as compared to healthy children, possibly indicating that autoantibodies to Endoglin are not the cause of tissue damage, but rather a consequence thereof (**Figure 7F**). Nevertheless, it is interesting to investigate a possible role for this protein, and antibodies targeting it, in the context of MIS-C and Kawasaki disease pathogensis.

**Figure 7.**
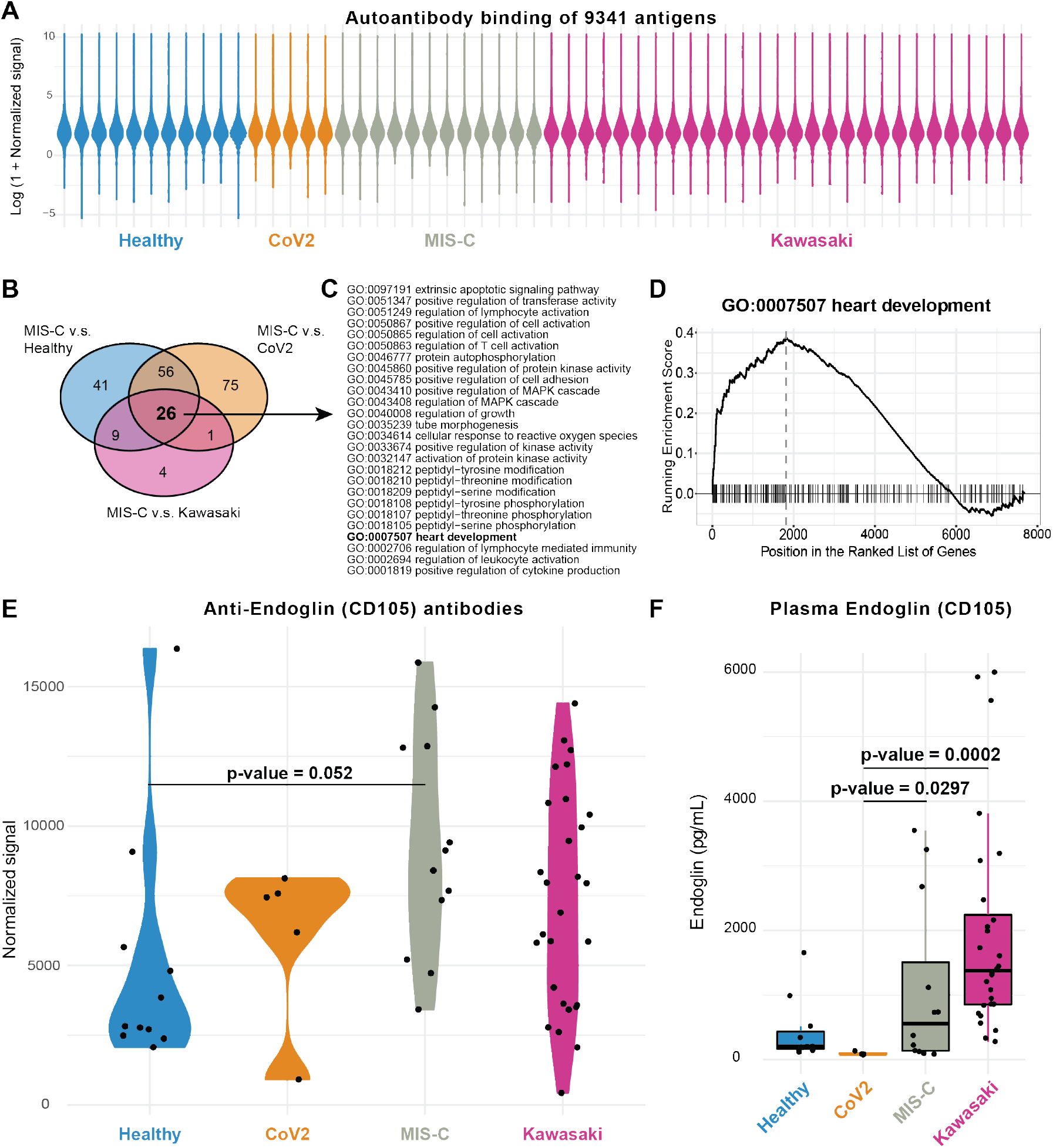
Autoantibodies in MIS-C, Kawasaki and healthy children. **(A)** Overall antibody binding intensities against 9341 antigens from 7669 human proteins in Healthy children (n=11), CoV2+ (n=5), MIS-C (n=12) and Kawasaki (n=28), violin plots colored by sample group. **(B)** Venn diagram showing 26 enriched GO-terms across MIS-C v.s. Healthy, Cov2+ and Kawasaki disease children. **(C)** The 26 GO terms enriched in MIS-C vs all other groups listed. **(D)** GSEA plot for G0:0007507 heart development. **(E)** Autoantibodies targeting the glycoprotein Endoglin (CD105). P-value comparing means in healthy children and MIS-C (FDR 1%). **(F)** Plasma Endoglin levels measured by ELISA in plasma samples.

Among the enriched GO-terms, the majority involve signal transduction and in particular signals mediated by phospho-tyrosine and -serine intermediates. GO: 0018100105 is one such term with strong overrepresentation among MIS-C samples over children with mild SARS-CoV-2 infection (Figure 8A). We found several possible autoantibodies in MIS-C-patients not seen in other samples such as antibodies to MAP2K2, and antibodies to three members of the Casein kinase family (**Figure 8B**). The latter is interesting in the context of SARS-CoV-2 infection because a global analysis of phosphorylation landscapes in cells infected by this virus showed strong upregulation of Casein-kinase 2 activity, and potent antiviral activity of its inhibitor drug silmitasertib (Bouhaddou et al., 2020). If this signaling cascade is important during viral infection, autoantibodies to intermediates of this pathway could be pathogenic downstream and give rise to MIS-C. These protein targets are also broadly expressed across tissues, which is in line with the diffuse clinical presentation in patients with MIS-C.

**Figure 8.**
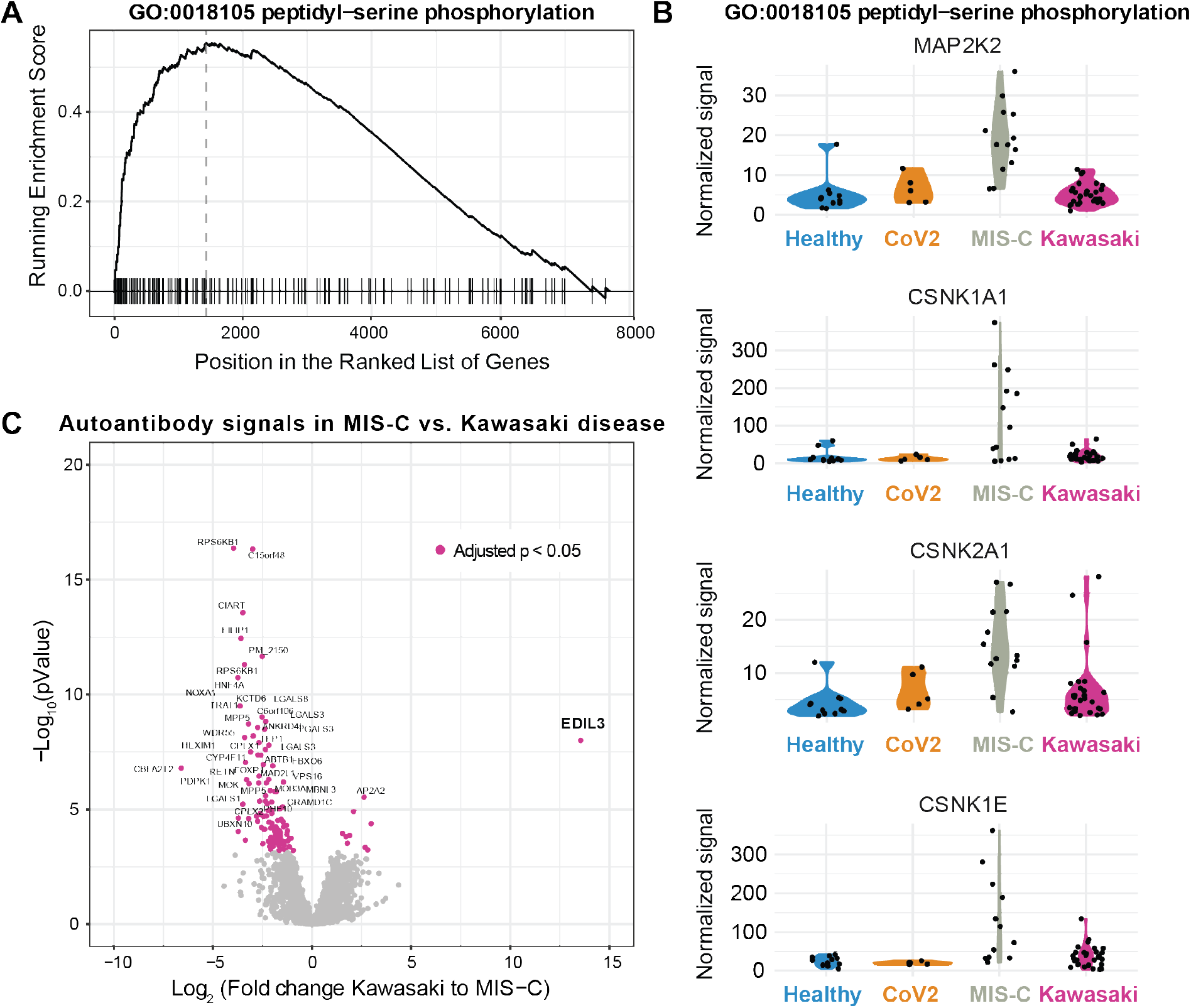
Candidate autoantibodies in MIS-C and Kawasaki disease. (A) GSEA ranked list of proteins belonging to the G0:0018105 term involving peptidyl-serine phophorylation. (B) Four candidate antigens bound by autoantibodies across the four patient groups but at highest levels in MIS-C children. **(C)** Volcano plot showing fold-change differences in autoantibody signals between Kawasaki disease (n=28) and MIS-C (n=13). Purple and annotated target antigens have p<0.05 (FDR 1%). EDIL3 is the single most overrepresented protein in Kawasaki disease.

Although there are overlapping features in the clinical presentation among MIS-C and Kawasaki disease patients, there are also clear differences. Kawasaki disease involves inflammation of medium sized arteries such as the coronary arteries, while MIS-C patients have more diffuse presentation involving the intestine, myocardium and brain. Also, we found that Kawasaki disease patients have higher levels of biomarkers associated with arterial damage and inflammation as compared to MIS-C patients. To understand possible differences in autoantibody profiles among these two conditions we focused on the autoantibodies most different between Kawasaki disease and MIS-C patients. Although there were a number of differentially regulated targets, EDIL3 autoantibodies were particularly abundant in Kawasaki disease patients but not in MIS-C (**Figure 8C**). This gene has previously been associated with Kawasaki disease (Jiang et al., 2017), and encodes a structural glycoprotein in the arterial vessel wall that is regulated by vascular injury. EDIL3 serves to inhibit inflammatory cell recruitment to the endothelium and when disrupted there is increased neutrophil recruitment to the tissue and elevated inflammatory responses (Choi et al., 2008). Collectively our results different cytokine profiles in Kawasaki disease and MIS-C, more biomarkers of arterial inflammation and damage in Kawasaki and a possible focus on autoantibodies to the Casein kinase system intermediates in MIS-C, a system strongly induced by SARS-CoV-2 infection. All these potential targets need to be validated in larger sets of MIS-C patients and more mechanistic experiments.

## DISCUSSION

The SARS-CoV-2 virus infects children at a similar rate as adults (Jones et al., 2020), but fortuitously the disease COVID-19 is very mild in the overwhelming majority of infected children (Brodin, 2020). Despite the mild COVID-19 in children, recent reports of severe hyperinflammatory disorders developing 1-2 months after the acute infection with SARS-CoV-2 are cause for much concern. This Multisystem Inflammatory Syndrome in Children associated with COVID-19 (MIS-C) also called Pediatric multisystem inflammatory syndrome, temporally associated with COVID-19 (PIMS-TS) have now been reported in more than 100 children worldwide and the numbers are increasing (Cheung et al., 2020; Moraleda et al., 2020; Riphagen et al., 2020; Verdoni et al., 2020; Whittaker et al., 2020).

Here we have combined high-dimensional analysis methods to uncover multiple aspects of the hyperinflammatory response in children with MIS-C. We find similarities with the immune response in Kawasaki disease, but also important differences, such as the IL-17A mediated hyperinflammation in Kawasaki disease, but not MIS-C. In addition, differences in T-cell subsets and cytokine mediators place MIS-C at the intersection of Kawasaki disease and acute SARS-CoV-2 infection immune states in children as well as the hyperinflammation seen in adults with severe COVID-19. We also find higher levels of biomarkers associated with arteritis and coronary artery disease in Kawasaki disease than in MIS-C suggesting a more diffuse endothelial involvement and immunopathology in MIS-C than in Kawasaki disease. Finally, we perform global autoantibody screening and find binding of autoantibodies to proteins involved in particular in immune cell signaling, structural proteins in heart and blood vessels. These data suggest possible targets of autoimmune attack. The hypothesis that autoantibodies contribute to the pathology in MIS-C is supported by the efficacy of intravenous immunoglobulin in MIS-C, a common approach to activate inhibitory Fc-receptors and prevent membrane-attack complexes by complement factors and thereby mitigating autoantibody-mediated pathology (Mackay et al., 2001).

Our results of lymphocyte subsets with lower naïve CD4^+^ T-cell, T_FH_ and increases in central and effector memory subpopulations in MIS-C as compared to Kawasaki disease patients could be partially explained by differences in patient age as immune cell proportions change with age even in healthy children (Olin et al., 2018; Schatorjé et al., 2012). We did however see these cell population differences remained even when comparing against additional older healthy controls, comparable in age to the MIS-C patients. Also, several studies have indicated defective T-cell responses as a central aspect of severe COVID-19 disease in adult patients (Vardhana and Wolchok, 2020). We find expanded CD57+ CD4^+^ T cells often representing immunosenescent and dysregulated T cells in both MIS-C and SARS-CoV-2+ children with mild infections, suggesting that this might be a direct consequence of the SARS-CoV-2 infection and a similarity between MIS-C and acute COVID-19. Additional studies in mild and especially asymptomatic COVID-19 cases are required to investigate this further.

Treatments of MIS-C have mostly followed protocols used in atypical Kawasaki disease given the overlapping presentation between these patients and children with MIS-C. The data presented here present a more complex picture with both shared features and clear differences that could influence strategies for treating these conditions. Kawasaki, but not MIS-C has a strong IL-17A upregulation and future clinical trials in severe Kawasaki disease patients treated with IL-17A blocking agents such as Secukinumab (Hueber et al., 2010) could be considered. In contrast the children in this study with MIS-C were treated with a combination of intravenous immunoglobulins (IVIG), corticosteroids and recombinant IL-1-receptor antagonist, IL-1RA (Anakinra). The profiles of seven cytokines significantly changed before and after treatment in patients with paired samples (**Figure 5B-C**). IVIG can neutralize some of the immunopathological effects of autoantibodies, while corticosteroids provide general immunosuppression and IL-1RA neutralizes the strong IL-1 response likely elicited by endothelial cells and innate immune cells recruited to the site of tissue injury. Other treatment strategies reported by others include TNFα blockade (Infliximab) (Whittaker et al., 2020), but compared to adults with acute COVID-19, TNFα levels are lower in MIS-C (**Figure 2**) and also when comparing TNFα-levels in MIS-C to that of healthy children we find no elevated levels (**Supplementary Figure 2B**), suggesting that TNFα-blockade will not be useful to mitigate MIS-C. Also, IL-6 blockade is a candidate treatment strategy already used in acute COVID-19 but we find highly variable I L-6 levels in MIS-C with 2/3 patients having normal I L-6 levels even during acute hyperinflammation (**Figure 4C)**. We therefore suspect that blocking I L-6 might not be universally effective in MIS-C patients. Overall the data presented here suggest novel directions for future work towards more mechanistic understanding of the immunopathology in MIS-C and development of novel immunomodulatory therapies that can mitigate the effects of hyperinflammatory disease in children during the COVID-19 pandemic.

## STAR METHODS

## CONTACT FOR REAGENT AND RESOURCE SHARING

Further information and requests for resources and reagents should be directed to Petter Brodin. Raw data is freely available for download: https://brodinlab.com/data-repository/

Scripts to reproduce the analyses are available through GitHub: https://github.com/Brodinlab/MIS-Cmanuscript

### Study Participants and Sample collection

45 SARS-CoV-2 infected children were enrolled within the CACTUS study at Bambino Gesù children hospital between March 17^th^ and May 15^th^ 2020. Diagnostic tests for SARS-CoV-2 infection are reported below. Three of the 45 Italian children found to be SARS-CoV-2+ by PCR herein were classified as having MIS-C. In addition, we enrolled 10 children at the Karolinska University Hospital fulfilling the same MIS-C WHO criteria (https://www.who.int/news-room/commentaries/detail/multisystem-inflammatory-syndrome-in-children-and-adolescents-with-covid-19). In summary this states that children and adolescents between 0 and 19 years old, with fever > 3 days, and displaying 2 signs among: i) rash or bilateral non purulent conjunctivitis or mucocutaneous inflammation signs, ii) hypotension or shock, iii) Features of myocardial dysfunction, pericarditis, valvulitis, or coronary abnormalities (including ECHO findings or elevated Troponin/NT-proBNP), iv) evidence of coagulopathy (by PT, PTT, elevated d-Dimers), v) acute gastrointestinal problems (diarrhea, vomiting, or abdominal pain); AND elevated markers of inflammation such as ESR, C-reactive protein, or procalcitonin; AND no other obvious microbial cause of inflammation, including bacterial sepsis, staphylococcal or streptococcal shock syndromes; AND evidence of SARS-CoV-2 infection (RT-PCR or serology positive), or likely exposure to COVID-19 patient. With regards to the MIS-C only 3 out of 4 patients were analyzed for deep immunology studies since one patient died due to cardiac complications after surgery during the study duration. Twenty-eight children diagnosed with Kawasaki disease were enrolled during their acute phase of the disease at the Academic Department of Pediatrics, Division of Immune and Infectious Diseases at Bambino Gesù Children’s Hospital (OPBG) between September 2017 and June 2019, well before the COVID-19 pandemic hit Europe. KD diagnosis was made according to the 2017 criteria of the American Heart association, including both the complete and incomplete types. Fever onset was considered as the first day of the acute KD phase. Twelve healthy controls (HC) were recruited at OPBG. Demographic and clinical data of all the patients are summarized in **Table 1**. Swedish patients are included in accordance with the Ethical permit Agency (Study ID: 2020-01911). Italian children are included within the study protocol approved by the OPBG Ethics Committees and informed consent was obtained from parents or guardians of all patients. Autoantibody analyses in APS-1 patients were approved by Ethical board of Stockholm (Permit: 2016/2553-31/2).

### Confirmation of SARS-CoV-2 infection

All patients enrolled in the CACTUS study with nasopharyngeal swab tested positive for SARS-CoV-2 nucleic acid using reverse-transcriptase qualitative PCR assay were considered confirmed cases of SARS-CoV-2 infection. In addition, all serum samples of COVID19+ were investigated for presence of SARS-CoV-2 antibodies. IgG antibodies were quantitatively tested by LIAISON® SARS-CoV-2 S1/S2 IgG test (Diasorin).

### Serum protein profiling and analyses

Serum proteins were analyzed using a multiplex technology based upon proximity-extension assays (Lundberg et al., 2011). We measured 180 unique proteins using the Olink panels of Immune Response and Inflammation (**Key Resource Table**). Briefly, each kit consisted of a microtiter plate for measuring 92 protein biomarkers in all 88 samples and each well contained 96 pairs of DNA-labeled antibody probes. To minimize inter- and intra-run variation, the data were normalized using both an internal control (extension control) and an inter-plate control, and then transformed using a pre-determined correction factor. The pre-processed data were provided in the arbitrary unit Normalized Protein Expression (NPX) on a log2 scale and where a high NPX represents high protein concentration.

Plasma protein analyses in **Figure 2** were performed using samples from children with MIS-C (n=13), children with Kawasaki (n=28), adult CoV2+ non-ICU (n=7 subjects, 14 time points total), and adult CoV2+ ICU (n=10 subjects, 43 time points total). In order to compare Adult CoV2+ cases with Kawasaki and MIS-C children, bridge normalization was used due to available bridge samples between assays regarding children. Bridge normalization was done with the function *olink_normalization()*. A 30% LOD cutoff was used to filter out proteins for all Olink plates. Then both plates were filtered for matching proteins across. The normalized combined plate with children cases as well as the plate containing the Adult CoV2+ cases were scaled within each set to unit variance (z-score) before merging. For the PCA and PC contribution plots, both were done using the library *factoextra* and only the top 20 contributions were displayed in the contribution plot.

Plasma protein analyses in **Figure 4** were performed using samples from healthy children (n=12), children with SARS-CoV-2 without hyperinflammation (n=41), children with MIS-C (n=21, pre-treatment n=11, post-treatment n=7), and children with Kawasaki (n=28). The four proteins (IL10, IL6, CCL11, IL5) that overlapped between Olink Immune Response and Inflammation panels were averaged. Proteins with >30% measurements below the threshold of detection were filtered out, resulting in 120 and 133 plasma proteins used for PCA in **Figures 2** and **4** respectively. Because Olink data from **Figure 4** was run in two batches, bridge normalization was applied using function olink_normalization from R package OlinkAnalyze, followed by batch correction using the function removeBatchEffect from R package limma. The analyzed Olink parameters for **Figures 2** and **4**, and their mean NPX and standard deviations are provided (**Supplementary Tables 1 and 2**). For analyses in **Figure 5**, seven paired pre/post treatment MIS-C samples were compared.

### VirScan

We investigated and compared antiviral antibody repertoires (IgG) of MIS-C, Cov2+, Kawasaki and healthy children with VirScan, a viral epitope scanning method that relies on bacteriophage display and immunoprecipitation. Briefly, VirScan uses a library of bacteriophages presenting 56-amino-acid-long linear peptides that overlap by 28 amino acids to collectively encompass the entire genomes of 1,276 viral strains from 206 viral species known to infect human cells. After virus inactivation and normalisation to total IgG concentration, plasma samples were incubated with the phage library to form IgG-phage immunocomplexes, later captured by magnetic beads. Collected bacteriophages were lysed and sequenced to identify the IgG-targeted epitopes. VirScore is the output given by VirScan and corresponds to the number of peptide hits that do not share epitopes, and it is incremented by 1 when a hit is enriched in the ‘output’ compared with the ‘input’. Background noise is usually observed in bacteriophage display assays. In order to minimise false-positive hits, VirScan removes hits occurring as a consequence of unspecific binding to beads only (blank reactions without antibodies). Moreover, it also removes cross-reactive antibodies by ignoring hits that share a subsequence of at least seven amino acids with any other enriched hit found in the same sample.

### Virscan data analysis

For each sample, sequencing reads were first mapped to the original library sequences with Bowtie, and the number of reads of each peptide in the original library were counted with SAMtools. A zero-inflated generalised Poisson distribution was applied to fit the frequency of peptides, and *-iog_l0_(P)* was calculated for each peptide as the probability of enrichment. Peptides with - *iog_l0_(P*) larger than 2.3 in both technical replicates were considered to be significantly enriched. Among the significantly enriched peptides, those appear in at least 3 of the beads samples were removed as nonspecific bindings. We also filtered out the peptides that were enriched in only one sample. The remaining enriched peptides were used to calculate the virus scores. To remove the hits caused by cross-reactive antibodies, we first sorted the virus by their total number of enriched peptides in descending order. For each virus in this order, we iterated through all the enriched peptides and removed those that shared a sequence of more than 7 amino acids with any previously observed peptides in any virus of the same sample. The remaining enriched peptides were considered specific and their number is the virus score for the virus. Afterwards, we filtered out the outlier viruses with Virus Scores >1 in only sample and negative Virus Scores in all others. We also filtered out viruses with Virus Scores < 2 in all samples.

### ELISA

Serum endoglin levels (CD105) were determined using manufacturer’s instruction (**Key Resource Table**). Briefly, samples were heat inactivated (56 C for 30 min) and ~50mL was used for ELISA-based quantification of Endoglin levels. Samples with out-of-range values were diluted and measured again

### Autoantibody profiling and analyses

We performed autoantibody profiling on samples from healthy children (n=11), children with CoV2+ (n=5), children with MIS-C (n=12) and children with Kawasaki (n=28) in two batches. Serum autoantibody reactivity was studied using full-length human protein arrays (ProtoArray v5.1, PAH05251020, ThermoFisher) (Zhu et al., 2001). Protein arrays were probed with serum at a dilution of 1:2000, and otherwise followed the protocol provided by the manufacturer for immune response biomarker profiling. Protein arrays were first incubated with blocking buffer (PA055, Life Technologies) for 1 hour, followed by 90 min incubation with serum at 1:2000 dilution, and 90 min incubation with detection antibodies: Alexa Fluor 647 goat anti-human IgG antibody (A21445, ThermoFisher) at 1:2000 dilution and Dylight 550 goat anti-GST (#DY550011-13-001, Cayman Chemicals) at 1:10,000 dilution. The LuxScan HT24 (BioCapital) microarray scanner was used. All analyses of autoantibody reactivities were performed using background-subtracted mean signal values of protein duplicates of the human IgG channel. Samples were quantile-normalized, and technical duplicates were averaged. For comparisons of autoantibody target reactivity between MIS-C and other groups (healthy control, CoV2+ and Kawasaki), differential expression was run using deseq function of the R package Deseq2. Ranked lists were obtained for each comparison to MIS-C (contrasts), and used for GSEA. GSEA was run on the ranked list of unique targets using the gseGO() function of the clusterProfiler R package.

### Flow Cytometry analysis of T cell subsets

Blood samples from KD and SARS-CoV-2+ patients were collected at the time of diagnosis and with regards to KD, always before Intravenous immunoglobulin administration. After ficoll, PBMCs and plasma samples were stored in liquid nitrogen or at -20°C, respectively, in Nunc Cryotubes (Merk KGaA, Darmstadt, Germany). Flow cytometry was performed for 30 patients, and isolated PBMCs were stained with LIVE/DEAD™ Fixable Near-IR Dead Cell Stain Kit (for 633 or 635 nm excitation, ThermoFisher, Waltham, Massachusetts, US) for 15 minutes at 4°C. Then, the cells were washed in wash buffer (phosphate-buffer saline with 1% bovine serum albumin) and stained for 30 minutes at 4°C with anti-hCD3 PE-CF594, anti-hCD4 BV510, anti-hCD25 PE, anti-hCD45RO-PerCP-Cy 5.5, anti-hCD27 V450, anti- hCD57 APC (all from BD Biosciences, Milan, Italy), anti-hCD127 PE-Cy7, anti-hPD1 BV711 (all from Biolegend, San Diego, CA). Data acquired by CytoFLEX cytometer (Beckman Coulter, Milan, Italy) were analyzed by FlowJo software v.10 (Treestar Software, Ashland, Oregon, USA).

### Flow cytometry data analysis

Statistical comparisons between two groups on flow cytometric frequencies were performed with t-test if both distributions were approximately normal or, conversely, with the Wilcoxon nonparametric test. In both cases, the normality of the distributions was tested with the D’Agostino-Pearson test. P-values less than 0.05 were considered to be statistically significant. Concerning clinical and routine laboratory data, median and interquartile range (IQ) for continuous variables were shown in **Table 1**. Continuous laboratory data (total blood count, CRP, AST, ALT, albumin, sodium, ferritin, triglycerides) and statistical comparison among different groups were performed as flow cytometric data analysis. Differences in gender distribution between two groups were tested with the Fisher test. MOFA was performed with the MOFA+ package (Argelaguet et al., 2019) (version 1.0) in the R statistical environment. Data analysis was performed using R (version 3.6.2).

## Data Availability

Raw data is freely available for download:
https://brodinlab.com/data-repository/
Scripts to reproduce the analyses are available through GitHub: https://github.com/Brodinlab/MIS-C_manuscript

## ACKNOWLEDGMENTS

We would like to acknowledge all patients and guardians who decided to participate to the study. We thank all the CACTUS study nurses team of the COVID-19 Center of “Bambino Gesù “Children’s Hospital” and Jennifer Faudella, Sonya Martin and Giulia Neccia for their precious administrative assistance. We thank SciLifeLab Plasma Profiling Facility for generating Olink data and the SciLifeLab autoimmunity profiling facility for instrument support.

## The CACTUS study team from the Children’s Hospital “Bambino Gesù”

Stefania Bernardi MD; Emma Concetta Manno MD; Paola Zangari MD; Lorenza Romani MD; Carlo Concato Bsc; Paola Pansa MD; Sara Chiurchiu MD; Andrea Finocchi MD,PhD; Caterina Cancrini Md,PhD; Laura Lancella MD; Laura Cursi MD; Maia De Luca MD; Renato Cutrera MD; Libera Sessa, PhD; Elena Morrocchi, PhD; Lorenza Putignani PhD; Francesca Calò Carducci MD; Maria A De Ioris MD; Patrizia D’Argenio MD; Marta Ciofi degli Atti MD; Carmen D’Amore MD.

